# AI-Enhanced Subtyping of Thymic Tumors: Attention-based MIL with Pathology-Specific Feature Extraction

**DOI:** 10.1101/2024.06.07.24308609

**Authors:** Haitham Kussaibi

## Abstract

**Purpose:** The precise classification of thymic tumors using whole slide images (WSIs) is essential for accurate diagnosis and treatment. While traditional Convolutional Neural Networks (CNNs) are commonly used for this purpose, emerging models tailored to pathology, such as Phikon and HistoEncoder, present promising alternatives as feature extractors. Additionally, the limited availability of annotated WSIs has driven the development of weakly-supervised classifiers like multiple-instance learning (MIL) models. In this study, we evaluate nine different combinations of extractor-classifier pairs for thymic tumor subtyping, including a novel, self-developed attention-based MIL classifier, AttenMIL.

**Methods:** The process began with curating a dataset of thymic tumor Whole Slide Images (WSIs) from the TCGA platform. Using the Yottixel method, patches were derived from these WSIs, and features were extracted from the patches using three different pathology-specific models: Phikon, HistoEncoder, and a pathology-fine-tuned ResNet50. The extracted features were then organized into small bags of instances through a chunking technique. Subsequently, three MIL classifiers AttenMIL, TransMIL, and Chowder were trained. Finally, the efficacy and generalizability of nine different combinations of extractor-classifier pairs were evaluated on unseen test images. Confusion matrices for each pair were utilized to provide insights into misclassification patterns and potential error sources.

**Results:** The Phikon feature extractor consistently delivered the highest classification accuracies, particularly when paired with the AttenMIL and Chowder classifiers, achieving up to 99% accuracy. This combination significantly outperformed other feature extractor-classifier pairs. Confusion matrices revealed that the AB and B3 subtypes were the most commonly confused classes across the different models.

**Conclusions:** The study demonstrates the potential of domain-specific feature extractors like Phikon, when coupled with robust MIL classifiers such as the novel AttenMIL and Chowder, in enhancing the accuracy and reliability of thymic tumor classification. The chunking-based augmentation method proved effective for thymic tumors, which are relatively homogeneous, but its applicability to heterogeneous tumors remains to be explored. Future research should address class imbalances and improve generalizability to different datasets.

**Statements and Declarations:** *Competing Interests:* The author has declared no competing interest. No funding was received for conducting this study.

*Declaration of generative AI and AI-assisted technologies in the writing process:* While preparing this work, the author used ChatGPT to improve language and readability. After using this tool/service, the author reviewed and edited the content as needed and took full responsibility for the publication’s content.

## 1 Introduction

Histopathology is the study of diseased tissues at the microscopic level, using various staining techniques and imaging modalities. It plays a crucial role in the diagnosis and classification of tumors, including thymic neoplasms.

Histopathological examination involves analyzing cellular morphology, and architectural patterns of tumor samples obtained through biopsy or surgical resection.

Thymic tumors, a type of anterior mediastinum neoplasm, originate from epithelial cells of the thymus gland. These tumors fall into two broad categories: thymomas and thymic carcinomas. Thymomas are typically slow-growing, while thymic carcinomas tend to be more aggressive, invasive, and prone to metastasizing. Thymomas display a diverse array of histological subtypes, formed by thymic epithelial cells with varying degrees of lymphocytic infiltration. The World Health Organization (WHO) classifies five types of thymomas based on their histological features: type A (spindle epithelial cell with rare lymphocytes), type AB (mixed A & B), type B1 (lymphocyte-rich), type B2 (equal epithelial and lymphocyte count), and type B3 (epithelial-rich) [1].

Accurate diagnosis and subtyping of thymic tumors are critical for determining appropriate treatment strategies and predicting patient prognosis.

However, this task can be challenging due to tumors’ complexity and variability, which include a broad spectrum of overlapping histological patterns and cellular compositions. Traditional diagnostic methods, such as microscopic examination of biopsy samples, are subjective and potentially susceptible to human error, interpretation biases, and inter-observer variability among pathologists. Consequently, there is a growing interest in producing more objective and reproducible tumor classification methods by leveraging the power of artificial intelligence (AI) and deep learning techniques.

Deep learning (DL), a subset of machine learning, employs artificial neural networks inspired by the structure and functionality of the human brain. These networks consist of multiple interconnected layers capable of learning hierarchical representations of data, allowing them to recognize complex patterns and make precise predictions. DL, particularly convolutional neural networks (CNNs), has demonstrated impressive performance in various medical imaging tasks in recent years [2]. CNNs are designed to extract relevant features from raw image data, making them well-suited for analyzing complex visual patterns and morphological characteristics present in histopathology images. These techniques have the potential to enhance diagnostic accuracy, reduce inter-observer variability, and provide quantitative analyses beneficial for clinical decision-making [2]. One challenge in applying CNN to histopathology images is the limited availability of large datasets necessary for training complex models. To address this limitation, researchers have implemented transfer learning techniques that adapt CNN models trained on a large dataset (such as ImageNet, a vast dataset of natural images) to a smaller, domain-specific dataset (such as histopathology images). This method allows the model to utilize the low-level features learned from the larger dataset while refining its higher layers to meet the specific task requirements [3]. However, the scarcity of annotations in histopathology Whole Slide Images (WSIs) necessitates the adoption of weakly-supervised approaches, such as multiple instance learning (MIL).

WSI, a huge histopathology image comprising hundreds of thousands of pixels, could generate hundreds or even thousands of small images/patches (called instances). The patches generated from each WSI are considered a single unit (or a bag of instances) sharing the same WSI label.

MIL models are designed to predict WSI/bag-level labels. Several well-established MIL models, including attention-based MIL, have proven effective in weakly-supervised medical image classification [4-6]. Herein, we introduce a simple, and robust attention-based MIL classifier (AttenMIL) to enhance the classification performance.

To address the limited number of WSIs in weakly-supervised tasks, we propose a novel method for dataset augmentation called ‘feature chunking’. In this method, WSI-derived features are split into smaller bags, each containing 200 features. This approach simulates the diagnostic process of pathologists, where a few relevant microscopic fields, rather than the entire slide, are sufficient for an accurate diagnosis.

On the other hand, unlike CNNs, which perform feature extraction and classification within the same model, training MILs require pre-extracted features using a separate model. Furthermore, while CNNs are effective at producing high-quality features, emerging pathology-specific models like Phikon [7] and HistoEncoder [8] are proving more efficient at extracting relevant and reliable features from histopathology images.

In the current study, we present a comprehensive analysis of AI-based subtyping of thymic tumors in WSIs. This analysis compares 3 domain-specific feature extractors combined with 3 MIL classifiers, in different combinations, describing end-to-end weakly-supervised WSI classification pipelines. In these pipelines, we employ the novel chunking method and introduce the novel AttenMIL classifier.

## 2 METHOD

The DL classification pipelines included several key stages: initial dataset preparation, model training, and performance evaluation (Fig. 1). This process began with the meticulous curation of datasets, starting with the collection of WSIs. Patches were then extracted from these images, and features were subsequently extracted from the patches using 3 different models. These extracted features were organized using chunking techniques. Three distinct classifiers were trained on the bagged features, and their performance was rigorously evaluated using unseen test images to assess their effectiveness and generalization capabilities.

**Fig. 1.**
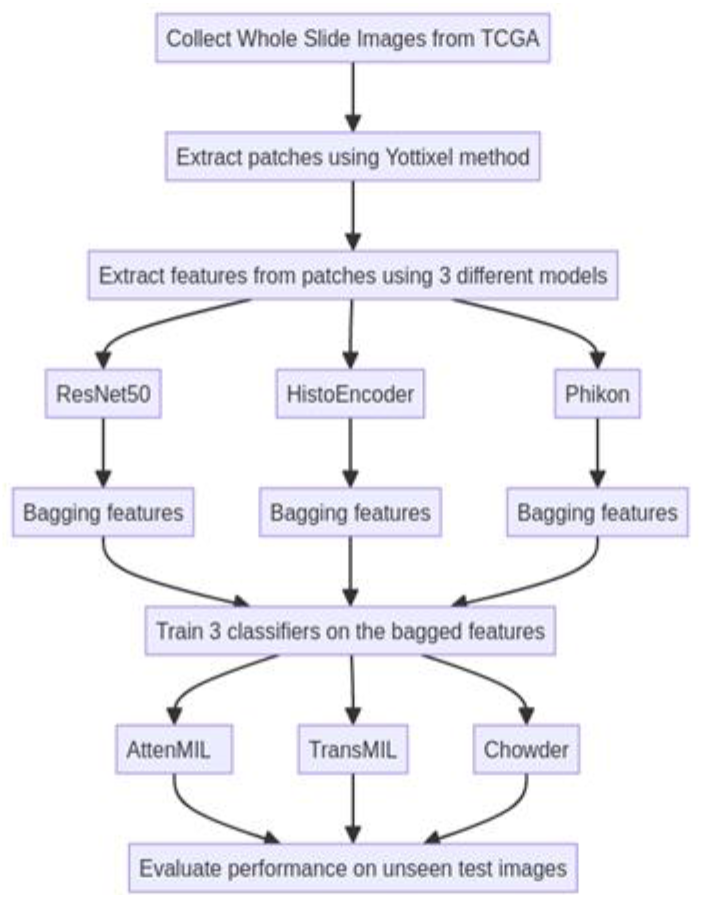
Flowchart illustrates the Overall Methodology (end-to-end pipeline), depicting the different stages and components involved in the AI-based classification process

### 2.1 Dataset Pre-processing

#### 2.1.1 Data Collection and Selection

The dataset comprised 242 histopathology WSIs of thymic epithelial tumors obtained from the publicly accessible TCGA database. These WSIs varied in size, ranging from 51 MB to 3.4 GB. The WSIs were categorized into six distinct subtypes/classes, with slide-level labels provided; detailed tumor annotations or masks were unavailable.

#### 2.1.2 Patch Extraction

WSIs are huge multilayer images with hundreds of thousands of pixels that necessitate being split into small patches to fit DL model training requirements.

To extract patches from tissue areas in WSIs, rather than blank backgrounds, we adopted a robust technique, called **Yottixel**, developed by KimiaLab [9]. Initially, Yottixel generates a tissue mask to isolate patches solely from tissue areas, maintaining a predetermined tissue-to-background ratio; patches with 30% background or more, were subsequently discarded. This method produces patches of size 250×250 pixels. Yottixel can also cluster the resulting patches, extract their features, and generate a barcode based on them to build an Image Search Engine. However, in this study, we only utilized the patch extraction part.

### 2.2 Feature Extraction

Before feature extraction, patches were standardized and resized to 224×224 pixels to comply with the requirements of extraction models.

During this phase, three distinct models were employed to extract features from patches:

#### ResNet50

A CNN pre-trained on ImageNet [10], and finetuned on the UBC-OCEAN dataset. This dataset contains more than 500 WSIs of different types of epithelial ovarian cancers [11-13].

#### Histoencoder

A pathology-specific model that has been trained in a self-supervised technique on Prostate cancer tissue images to extract and cluster features from histology images [8].

#### Phikon

Another self-supervised learning model tailored to extract features from histology images [7].

Phikon has its preprocessing function (AutoImageProcessor), while the other 2 models needed a custom normalization technique.

### 2.3 Construction of Features Bags

Instead of treating the features extracted from each WSI as a unit/bag, and consequently getting a limited number of bags for training, a pioneering chunks-based method was introduced. This innovative approach involves grouping features into bags of uniform size, each containing 200 features/instances. This process is achieved by the (torch.chunk) function. Employing this technique not only increased the size of the training dataset by a factor of ten (as shown in Table 1) but also produced bags of consistent sizes, eliminating the need for additional padding measures.

**Table 1:** Dataset distribution among thymic tumor subtypes.

| Tumors' Subtypes | A | AB | B1 | B2 | B3 | CA |
| --- | --- | --- | --- | --- | --- | --- |
| N. of WSIs | 34 | 68 | 39 | 61 | 22 | 18 |
| N. of Patches/Thousand | 56 | 120 | 80 | 102 | 40 | 30 |
| N. of Bags (200-sized) | 280 | 600 | 400 | 510 | 200 | 150 |

Although, this method works very well in thymic tumors, which are relatively homogeneous; however, in heterogeneous tumors, larger-sized bags or an additional tumor segmentation phase might be required.

The following table presents the dataset with the distribution of thymic tumor subtypes including the number of WSIs, the patches/features extracted, and the corresponding number of bags created.

### 2.4 Classification Models

Given the weakly supervised nature of our approach, which involves slide/bag-level labels, the most suitable classifiers were those based on Multiple Instance Learning (MIL). In MIL, the training data is organized into bags, with each bag containing multiple instances that share a single label. In this study, we experimented with two well-known MIL models (TransMIL and Chowder), as well as a novel self-built, attention-based MIL classifier named ‘AttenMIL’.

*AttenMIL* incorporates attention mechanisms within a MIL architecture. The attention mechanism allows the model to focus on specific regions or features within the input data, enhancing its ability to capture relevant information. This model is particularly effective in tasks where certain parts of input data are more informative than others. AttenMIL is composed of the following layers:

1. *Normalization function* ensures that the input data is on a consistent scale, which is crucial for the stability and performance of neural network models. 2. *Instance Normalization Layer* normalizes the input data instance-wise, improving the model’s robustness. 3. *Fully Connected Layers* with ReLU activations and dropout for regularization. 4. *Attention Mechanism* computes attention scores for instance-level predictions, allowing the model to focus on the most relevant parts of the input. 5. *Bag-level prediction* uses attention scores to aggregate instance-level predictions into a final bag-level output.

*TransMil* (Transformer based Correlated Multiple Instance Learning for WSI Classification) [5]: Unlike traditional MIL methods that treat instances independently, TransMIL accounts for the correlation between different instances within a WSI. This approach uses a transformer model to capture both morphological and spatial information from images.

*Chowder* [14]: A MIL-based classifier designed for weakly supervised learning tasks. It employs a combination of convolutional neural networks (CNNs) and MIL techniques to learn discriminative representations from bag-level labels. This involves embedding the features from individual image tiles and aggregating the most relevant positive and negative evidence tiles to make predictions.

These 3 classifiers are specifically tailored for tasks where only limited labeling is available, making them suitable choices for scenarios where annotations are provided at a higher level of granularity, such as slide/bag-level labels.

### 2.5 Classifier Training & Evaluation

#### 2.5.1 Experimental Setup

##### Train-test Split

The dataset was partitioned into training and testing sets using an 80% train and 20% test split, ensuring a stratified distribution across classes. This approach maintained a proportional representation of each tumor subtype in both training and testing subsets, thus minimizing bias in model evaluation.

##### Hardware Specifications

The experiments were conducted on a robust hardware setup comprising a GPU with 12 GB of memory and 8000 CUDA cores, supplemented by 32 GB of RAM and an Intel Core i7 13th Generation processor. This high-performance configuration enabled efficient training and evaluation of the deep learning models.

##### Implementation Framework

PyTorch, a widely adopted deep learning framework renowned for its flexibility and scalability, served as the primary implementation tool. Its intuitive interface and extensive library of pre-built modules facilitated seamless development and experimentation with complex neural network architectures.

##### Hyperparameters

Several key hyperparameters were meticulously tuned to optimize model performance. The batch size was set to 32, striking a balance between computational efficiency and model convergence. Number of epochs was set at 100. We utilized Adams as an optimizer with a learning rate scheduler (the initial learning rate was configured to start at 0.001). Additional hyperparameters fine-tuning tailored to each specific model architecture and training regimen was conducted iteratively in cross-validation training loops to maximize model efficacy and generalization capabilities.

#### 2.5.2 Training Process

The training process involved training these deep learning architectures to effectively classify the features extracted in the preceding step and associate them with their respective tumor subtypes. This encompassed two key phases: training and validation, where meticulous attention was paid to prevent over-fitting and enhance model generalization.

Despite the relatively high number of epochs set at 100, a cautious approach was taken by incorporating early stopping with a patience of 10 into the training regimen. This strategic measure ensured that training ceased when the model’s performance plateaued on the validation set, thus mitigating the risk of overfitting. By monitoring the model’s performance at regular intervals during training, early stopping provided a safeguard against the potential deleterious effects of over-training, allowing the model to attain optimal performance while maintaining generalization capabilities.

The decision to employ the Adam optimizer stemmed from its proven efficacy in iteratively updating the model’s parameters to minimize classification error. To further enhance its performance, the Adam optimizer was integrated with a learning rate scheduling mechanism with patience of 3. This adaptive technique dynamically modulated the learning rate throughout the training process, thereby facilitating smoother convergence toward the optimal solution. By intelligently adjusting the learning rate based on the observed performance, the model could navigate through the optimization landscape more efficiently, ultimately enhancing its convergence speed and overall effectiveness.

Addressing the variability in class sizes, a customized loss function named **ClassWeightedCrossEntropyLoss** was utilized. This loss function assigned higher weights to less frequent classes, thereby alleviating the impact of class imbalances on model training.

Furthermore, to comprehensively assess model performance and robustness, a cross-validation strategy employing 5 K-folds with an 80/20 split ratio and 32 batch size, was implemented. This technique partitioned the dataset into five equally sized folds, iteratively training the model on four folds and validating on the remaining fold to obtain reliable estimates of performance metrics.

#### 2.5.3 Classifier Inference Procedure

To evaluate the performance of each combination extractor-classifier on an unseen test dataset, the best-performing weight, across all the five folds of the cross-validation training process, has been chosen.

#### 2.5.4 Performance Metrics

In evaluating the efficacy of classification models for thymic tumors, a comprehensive suite of evaluation metrics has been utilized. These metrics serve as quantitative measures by which the model’s predictions are compared against the ground truth labels provided by expert pathologists (dataset labels).

Evaluation metrics employed in this study include:

##### Accuracy

represents the overall proportion of correctly classified samples across all classes. It provides a broad assessment of the model’s performance in accurately predicting tumor subtypes.

##### Confusion Matrix

A fundamental tool in performance evaluation, the confusion matrix provides a detailed breakdown of correct and incorrect classifications for each class. It offers insights into the model’s strengths and weaknesses, facilitating a granular analysis of its predictive capabilities across different tumor subtypes.

## 3 Results

### 3.1 Extracted features

The following table provides valuable information regarding the dimensions (number of features per patch) and storage requirements of the extracted features (Table 2).

**Table 2:** Extracted features by each DL model.

| Extractor | ResNet50 | Histoencoder | Phikon |
| --- | --- | --- | --- |
| N. of Features | 2048 | 1152 | 768 |
| Required Storage | 4.4 GB | 2.5 GB | 1.6 GB |

### 3.2 Performance metrics

Best Overall Performance: The combination of Phikon as a feature extractor with either AttenMIL or Chowder as classifiers demonstrated the highest performance with near-perfect scores.

The classification accuracies of the different combinations (extractor-classifier) are shown in (Fig. 2).

**Fig. 2.**
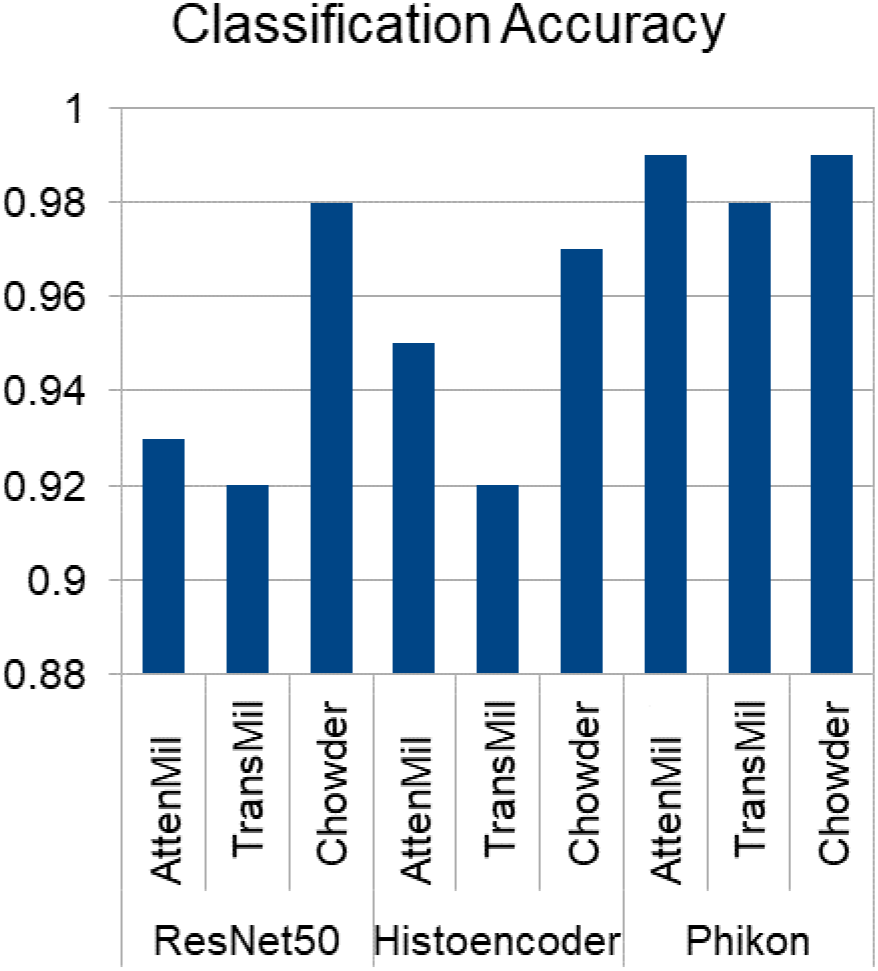
Classification accuracy of the different combinations (extractor-classifier)

### 3.3 Confusion Matrix and Error Analysis

The confusion matrices for each feature extractor-classifier pair provide insights into misclassification patterns and potential error sources (table 3).

**Table 3:** Accuracy of Thymic Tumor prediction by Different Extractor-Classifier Combinations for each subtype.

| Class | R-A | R-T | R-C | H-A | H-T | H-C | P-A | P-T | P-C | AvgClass |
| --- | --- | --- | --- | --- | --- | --- | --- | --- | --- | --- |
| A | 100.0% | 100.0% | 100.0% | 87.5% | 92.8% | 96.4% | 98.5% | 100.0% | 100.0% | 97.24% |
| AB | 88.7% | 93.2% | 97.8% | 97.4% | 94.9% | 97.5% | 98.2% | 97.4% | 98.1% | 95.91% |
| B1 | 84.8% | 88.5% | 98.1% | 94.4% | 87.6% | 96.4% | 98.9% | 98.8% | 100.0% | 94.17% |
| B2 | 97.8% | 88.5% | 99.3% | 96.9% | 92.1% | 96.0% | 99.2% | 97.6% | 99.2% | 96.29% |
| B3 | 95.7% | 92.1% | 93.6% | 97.9% | 88.0% | 97.9% | 97.8% | 92.7% | 97.7% | 94.82% |
| CA | 97.1% | 93.1% | 100.0% | 90.9% | 97.7% | 100.0% | 100.0% | 97.3% | 100.0% | 97.34% |
| AvgCom | 94.02% | 92.57% | 98.13% | 94.17% | 92.18% | 97.37% | 98.77% | 97.30% | 99.17% |  |

R: ResNet50, H: HistoEncoder, P: Phikon, A: AttenMIL, T: TransMil, C: Chowder AvgCom: Average accuracy for each combination across all classes. AvgClass: Average accuracy for each class across all combinations.

*ResNet50***-***AttenMIL* model performs very well in predicting class A, B2, and CA, with 100%, 97.8%, and 97.1% accuracy respectively. It also does well on class B3, correctly predicting 95.7% of samples with 4.4% of B3 predicted as AB and B2. The model struggles most with classes AB (88.7% correct), and B1 (84.8%) with 8.7% of AB and 10.1% of B1 samples misclassified as B2.

*ResNet50***-***TransMil* also excels at predicting class A (100% accuracy). It performs well on class AB (93.2), CA (93.1%), and B3 (92.1%), while it struggles with class B1 (88.5%) and B2 (88.5%). There is some confusion between B1 and B2 (6.9% of B1 is misclassified as B2 and 4.3% of B2 is misclassified as B1).

*ResNet50***-***Chowder* performs exceptionally well on class A and CA, with 100% accuracy for both. It also does very well on classes B2 (99.3%), B1 (98.1%), and AB (97.8%). The main issue was with class B3 (93.6%) with 4.3% confused as AB.

*HistoEncoder***-***AttenMIL* achieves high accuracy for classes B3 (97.9%) and AB (97.4%). Performance is also strong for class B2 at 96.9% and Class B1 at 94.4%. However, it struggles with classes A, and CA, with accuracies of 87.5%, and 90.9% respectively. There is notable confusion between classes A with AB and B3. Furthermore, a high percentage (9.1%) of CA is weirdly misclassified as B3.

*HistoEncoder-TransMil* delivers the best performance on class CA at 97.7% accuracy. It also performs well on classes AB (94.9%), A (92.8%), and B2 (92.1%). However, there is more confusion with classes B1 (87.6%) and B3 (88.0%), with 8% of B3 cases misclassified as class AB.

*HistoEncoder-Chowder* performs extremely well for class CA, achieving 100% accuracy. It also does well on classes B3 (97.9%), and AB (97.5%). Performance on classes B1, A, and B2 is slightly lower at (96.4%, 96.4%, and 96.0% respectively). There was 6% confusion between B1 and B2 cases.

*Phikon-AttenMIL* (**Fig. 3**) performs very well at classifying class CA, with 100% accuracy. It also does well in classes A, AB, B1, and B2 with 98.5%, 98.2%, 98.9%, and 99.2% accuracy respectively. Performance is slightly lower on class B3 at 97.8% accuracy. There is very little confusion between the classes, 2.2% of B3 cases are misclassified as B2, and 1.5% of A cases are misclassified as AB.

**Fig. 3.**
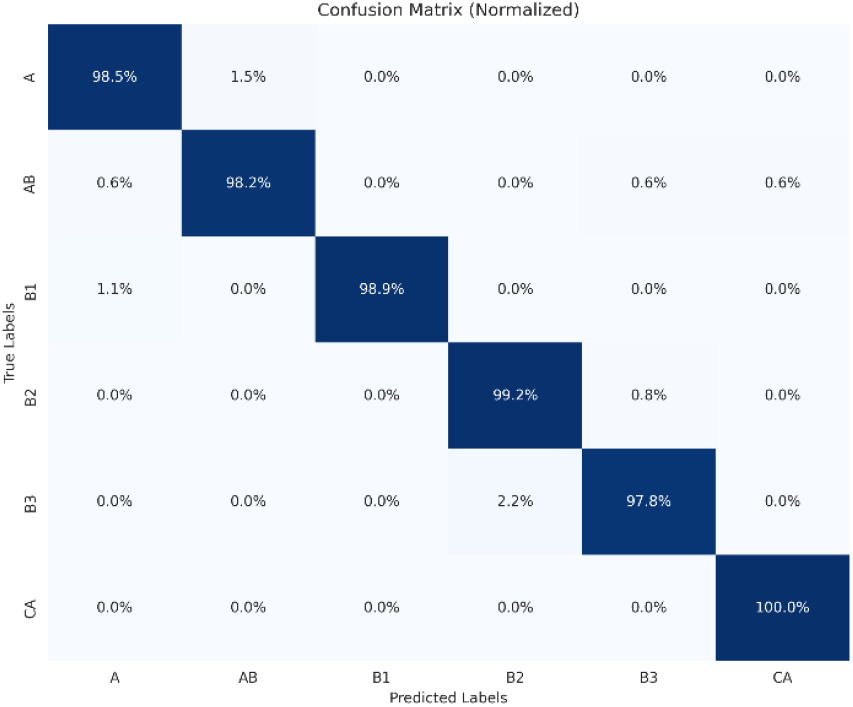
Confusion matrix for the Phikon-AttenMIL model

*Phikon-TransMil* has 100% accuracy on class A. It performs well on classes AB, B1, B2, and CA at 97.4%, 98.8%, 97.6%, and 97.3% respectively.

Accuracy is lower on class B3 at 92.7%. There is more confusion compared to the other two classifiers. A 7.6% of B3 cases are misclassified as AB and B2.

*Phikon-Chowder* (**Fig. 4**) achieves 100% accuracy on classes A, B1, and CA. Performance is very high on classes AB, B2, and B3 at 98.1%, 99.2%, and 97.7% respectively. There is slight confusion in some classes with 2.3% of B3 cases misclassified as AB, and 1.3% of AB cases misclassified as B2.

**Fig. 4.**
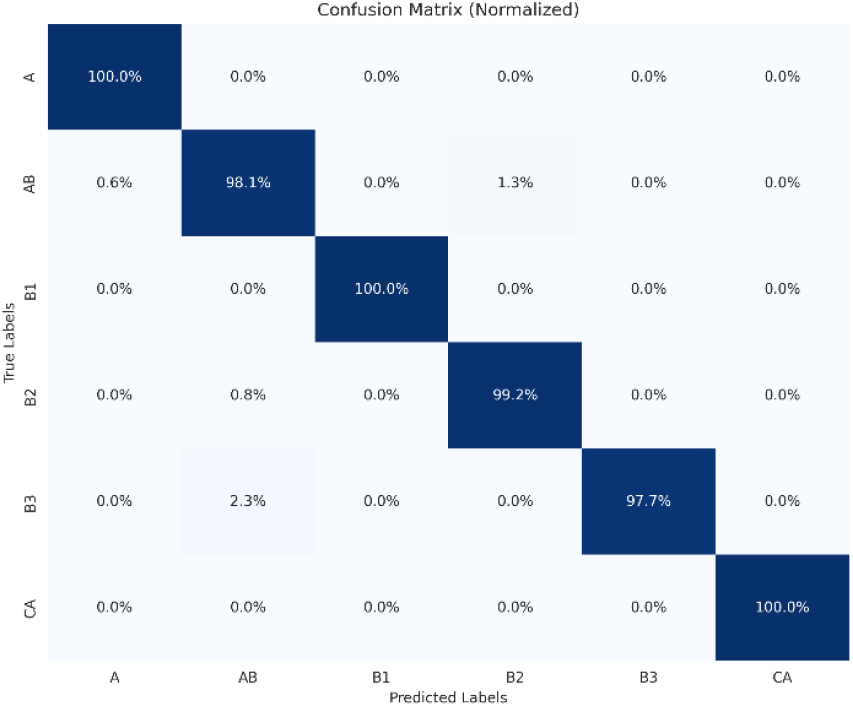
Confusion matrix for the Phikon-Chowder model

## 4 Discussion

In this study, we evaluated the performance of nine different deep-learning pipelines for subtyping thymic tumors WSIs into six classes (A, AB, B1, B2, B3, CA). The pipelines were constructed by combining a pathology-specific feature extractor (ResNet50, HistoEncoder, or Phikon) with a MIL classifier (AttenMIL, TransMil, or Chowder).

### 4.1 Analysis of Classification Performance

The classification performance of the proposed models is evaluated using accuracy and confusion matrices. By rigorously analyzing these metrics, we could gain insights into the strengths and limitations of these models, allowing for informed comparisons between approaches.

The study reports impressive results, with most combinations achieving accuracies above 0.92 and some even reaching near-perfect accuracy (0.98-0.99). Our results show that the choice of feature extractor and classifier had a significant impact on the overall model performance.

ResNet50, a well-established architecture, demonstrated high accuracy with Chowder (0.98) but lower accuracy with the other 2 classifiers (0.93, 0.92). Overall, its accuracy across all classifiers was above 0.92 which is better than a previous study that reported (AUC 0.91 and 0.9) using ResNet50-Chowder for lung tumors and lymph node metastasis classification tasks respectively [14]. However, the model was only pre-trained on ImageNet in that study, while our version was fine-tuned on histopathology images.

ResNet50’s primary weakness lies in its computational complexity and resource-intensive nature. Extracted features are of high dimensionality, leading to increased storage requirements and computational overhead during training and inference.

Histoencoder, a self-supervised learning foundation model designed to extract features from histopathology images, showed competitive performance, especially with AttenMIL and Chowder (Acc. 0.95 and 0.97). This is also better than a recent study exploring the potential of Histoencoder to classify breast cancers (Accuracy 0.89) [15]. While its average performance across all the classifiers is close to ResNet50, the smaller size of extracted features (1152) compared to ResNet50 (2048) may indicate a potential trade-off between feature richness and classification performance. The current available Histoencoder is pre-trained on prostate cancer images which are different in morphology and architecture from thymoma, this might explain its modest achievement, however, Histoencoder proved its superiority in features clustering and tissue segmentation [8].

Phikon, another histopathology-specific self-supervised learning model, exhibits outstanding results across the board, with near-perfect metrics (0.98-0.99) for all classifiers. Phikon’s ability to learn discriminative representations from histopathological data contributes to its exceptional performance. This novel approach advances the field by demonstrating the effectiveness of domain-specific self-supervised learning in capturing discriminative features for thymic tumor classification. Its extracted features, while useful, are smaller in size compared (768) to ResNet50, and HistoEncoder, possibly indicating a trade-off between feature richness and performance. However, its exceptionally high-performance warrants further validation on external datasets to assess generalization and rule out overfitting.

The introduction of self-supervised domain-specific learning models, especially Phikon, represents a notable improvement over the traditional CNN-based approach. These models demonstrate superior performance in tumor classification tasks, underscoring the importance of leveraging domain-specific features for enhanced accuracy and robustness.

On the other hand, using MIL classifiers in conjunction with domain-specific feature extractors further enhances performance by effectively utilizing bag-level labels in weakly supervised settings.

AttenMIL, while simple, showed impressive results when combined with the proper domain-specific feature extractor (Phikon).

Through careful hyperparameter tuning, regularization techniques, and model selection, we have optimized the performance of each approach, achieving commendable accuracy scores across different combinations of extractors and classifiers.

### 4.3 Confusion Matrix and Error Analysis

The confusion matrices for each feature extractor-classifier pair provide insights into misclassification patterns and potential error sources. All the nine confusion matrices corresponding to the nine different combinations are available on the current project repository: https://github.com/hkussaibi.

The Phikon extractor consistently yielded the highest accuracies when paired with any of the three classifiers, with most class-level accuracies exceeding 97%. The HistoEncoder and ResNet50 extractors also performed well but had slightly lower accuracies and more inter-class confusion compared to Phikon.

Regarding classifiers, AttenMIL and Chowder generally outperformed TransMil when combined with any feature extractors. AttenMIL demonstrated the best overall performance, achieving the lowest inter-class confusion. Chowder delivered comparable results with slightly more confusion between certain classes. TransMil had lower accuracies on some classes and more misclassifications compared to the other two classifiers.

These trends suggest that Phikon paired with Chowder or AttenMIL offers the most robust performance for thymic tumor classification, while ResNet50 and HistoEncoder also perform well but with specific areas of improvement needed.

Based on these findings, we recommend using the Phikon extractor combined with either AttenMIL or Chowder classifier for thymic tumor classification tasks. These combinations delivered the best performance in our experiments. However, the specific choice may depend on the relative importance of accuracy versus inter-class confusion for a given use case.

Achieving high accuracy for class A, and CA is a notable strength across all combinations, indicating a relatively easier classification for this tumor subtype due to its distinct morphology from other classes.

The models struggled in correctly predicting certain types like B3 which might be explained by the low number of B3 training cases and also by a relatively close morphology with other types.

To further improve classification performance, we suggest focusing on better-differentiating types AB, B1, and B3, as those were the most commonly confused classes. Collecting more training data, especially for the rare types like B3, may help the models learn to distinguish these challenging cases. Experimenting with other neural network architectures and hyperparameter settings could also potentially boost performance.

### 4.4 Potential Biases and Limitations

While the current study demonstrates significant progress in applying deep learning techniques to the classification of thymic tumors, it is important to acknowledge potential biases and limitations in the dataset, methodology, and results.

Class imbalances, where certain thymic tumor subtypes may be under-represented, can lead to biases and reduced performance for those classes.

Thymic tumors are relatively homogeneous, which explains the high performance of chunking-based augmentation of the dataset. However, this method might not work well with heterogeneous tumors, or WSIs that include only a focal tumor area. In such cases, using larger-sized bags or incorporating a pre-segmentation step is necessary.

The generalizability of the models to unseen data from different sources or acquired under different conditions should also be considered.

Histopathology images can be affected by various artifacts, such as staining variations, tissue folds, or image quality issues, which may impact the model’s performance. Ensuring robustness to these artifacts and noise is crucial for reliable classification results.

On the other hand, the interpretability of pathology-specific self-supervised models (Histoencoder & Phikon) remains a concern, as they are often perceived as “black boxes” making it difficult to understand the reasoning behind their predictions. This lack of transparency can hinder trust and acceptance among healthcare professionals.

### 4.5 Future Research Directions

Future work could include incorporating additional data augmentation techniques to address class imbalances, improve generalization, and enhance clinical trust and adoption. Acquiring larger, high-quality, and diverse datasets through collaborative efforts among multiple medical centers and standardized data collection protocols can improve the generalization and robustness of the models. Collaborations between AI experts and pathologists to validate findings with prospective clinical validation studies to assess real-world performance will be crucial for translating these research advances into clinical practice and integrating them into diagnostic workflows.

Exploring multi-modal data fusion, such as incorporating radiology images, genetic information, or electronic health records, could potentially improve the accuracy and comprehensiveness of thymic tumor classification. Integrating complementary information from different data sources may provide a more holistic understanding of the disease.

Developing transparent and explainable AI systems can enhance trust among healthcare professionals and facilitate accountability.

Finally, as new data becomes available and our understanding of thymic tumors evolves, developing frameworks for continuously updating and improving AI-based classification models will be necessary. This ensures that the models remain up-to-date and incorporate the latest knowledge and advancements in the field.

## 5 Conclusion

In conclusion, our study demonstrates that deep learning models can accurately classify thymic tumor histology images, but the choice of feature extractor and classifier significantly impacts performance. The Phikon extractor combined with AttenMIL or Chowder appears to be the most promising approach. With further refinements, these models could be valuable tools for assisting pathologists and informing thymic tumor treatment decisions.

## Data Availability

All data produced in the present study are available upon reasonable request to the author

https://github.com/hkussaibi

## Acknowledgment

The results shown here are based on data generated by the TCGA Research Network: https://www.cancer.gov/tcga.

## Code availability

The full pipelines will be available on (https://github.com/hkussaibi).

